# Reorganization of Substance Use Treatment and Harm Reduction Services during the COVID-19 Pandemic: A Global Survey

**DOI:** 10.1101/2020.09.21.20199133

**Authors:** Seyed Ramin Radfar, Cornelis A J De Jong, Ali Farhoudian, Mohsen Ebrahimi, Parnian Rafei, Mehrnoosh Vahidi, Masud Yunesian, Christos Kouimtsidis, Shalini Arunogiri, Omid Massah, Abbas Deylamizadeh, Kathleen T Brady, Anja Busse

## Abstract

**Background:** The COVID-19 pandemic has impacted people with substance use disorders (SUDs) worldwide and healthcare systems have reorganized their services in response to the pandemic.

**Methods:** One week after the announcement of the COVID-19 as a pandemic, in a global survey, 177 addiction medicine professionals described COVID-19-related health responses in their own 77 countries in terms of SUD treatment and harm reduction services. The health response is categorized around (1) managerial measures and systems, (2) logistics, (3) service providers and (4) vulnerable groups.

**Results:** Respondents from over 88% of countries reported that core medical and psychiatric care for SUDs had continued; however, only 56% of countries reported having had any business continuity plan, and, 37.5% of countries reported shortages of methadone or buprenorphine supplies. Participants of 41% of countries reported partial discontinuation of harm-reduction services such as needle and syringe programs and condom distribution. 57% of overdose prevention interventions and 81% of outreach services also having been negatively impacted.

**Conclusions:** Participants reported that SUD treatment and harm reduction services had been significantly impacted globally early during the COVID-19 pandemic. Based on our findings, we provide a series of recommendations to support countries to be prepared more efficiently for future waves or similar pandemics to 1) help policymakers generate business continuity plans, 2) maintain use of evidence-based interventions for people with SUDs, 3) be prepared for adequate medication supplies, 4) integrate harm reduction programs with other treatment modalities and 5) have specific considerations for vulnerable groups such as immigrants and refugees.

**Highlights:**

- COVID-19 negatively impacted services for PWSUD globally.
- Addiction medicine downgraded more than other psychiatry services.
- Business continuity plan for PWSUD services reported only in about half of the countries.
- Refugees & migrants had more negative impact compared to other vulnerable groups.
- Harm reduction services discontinued partially or totally during pandemic.

## 1. Introduction

Coronavirus disease 2019 (COVID-19) was announced as a pandemic by the World Health Organization (WHO) on March 11th, 2020 (“Coronavirus disease (COVID-19) pandemic,” 2020). COVID-19 quickly became a global concern given the rapid transmission of SARS-CoV-2 (the infectious agent), lack of a vaccine or evidence-based treatments, person-to-person airborne spread of SARS-CoV-2 and high mortality of COVID-19 in specific populations, especially marginalized groups and/or those with pre-existing conditions (Han, Kim, Chung, Park, & Cheong, 2018; Masozera, Bailey, & Kerchner, 2007; Onder, Rezza, & Brusaferro, 2020). Lack of capacity to anticipate, cope with, resist, and recover from COVID-19-related health consequences are related to individual vulnerability (Adger, 1999). To manage the current situation as best as possible, vulnerable groups should be recognized and helped with special considerations by relevant health systems (Marsden et al., 2020).

According to the World Drug Report 2020, among approximately 269 million people with past-year drug use, over 35 million people experienced substance use disorders (SUDs) (Knopf, 2020). People with SUDs (PWSUDs) may be particularly vulnerable to COVID-19 and complications for multiple reasons (Volkow, 2020). PWSUDs experience underlying diseases that constitute risk factors for COVID-19 infection or can be exacerbated by it; for instance, long-term use of substances may cause cardiovascular problems (Havakuk, Rezkalla, & Kloner, 2017) and chronic obstructive pulmonary disease (Riezzo et al., 2012). Such co-morbidities may exacerbate superimposed COVID-19 symptoms and lead to higher mortality rates (Arya & Gupta, 2020; Lai, Shih, Ko, Tang, & Hsueh, 2020). Poor immune system functioning is also prevalent in PWSUDs as a result of chronic alcohol and drug use and blood-borne or sexually transmitted illnesses (Cook, 1998; Szabo & Mandrekar, 2009), poor nutritional status (Bhaskaram, 2002), and socioeconomic factors (Spooner & Hetherington, 2005). Among PWSUDs, people who inject drugs (PWIDs) are at particularly high risk of COVID-19, as well as overdoses, unsafe injections and risky sex (Vasylyeva, Smyrnov, Strathdee, & Friedman, 2020).

Psychological conditions (e.g. phobia, anxiety and panic attacks) during natural disasters and pandemics that may precipitated, perpetuated or exacerbated through social isolation and quarantine, may lead at-risk people to start and/or relapse into drug-taking (Arya & Gupta, 2020; Nobles, Martin, Dawson, Moran, & Savovic, 2020). Psychiatric comorbidity has a negative impact on recovery from COVID-19 and may increase risk of non-fatal and fatal overdoses and suicides (Clay & Parker, 2020; De Sousa, Mohandas, & Javed, 2020; Frank, Fatke, Frank, Förstl, & Hölzle, 2020; Nobles et al., 2020). In the general population, COVID-19 and related concerns such as potential mortality may act as internal stressors (Liu & Doan, 2020) and promote cognitive impairments (Zarrabian & Hassani-Abharian, 2020) in domains such as decision-making (Starcke & Brand, 2012), problem-solving (Cheng & Lam, 1997), and attention (Dutra, Marx, McGlinchey, DeGutis, & Esterman, 2018), and thus may increase the incidence and prevalence of psychiatric disorders including PWSUDs (Fiorillo & Gorwood, 2020; Pfefferbaum & North, 2020).

Stigma may undermine social cohesion, contributing to situations in which the virus is more, not less, likely to spread. Such spread may result in more severe health problems and difficulties controlling a disease outbreak (Ren, Gao, & Chen, 2020). There is an elevated likelihood for PWSUDs to be homeless and live in crowded shelters and neighborhoods (Coetzee & Kagee, 2020). Synergistically, poor economic status linked to limited accessibility to health care (Ahern, Stuber, & Galea, 2007; O’Sullivan & Bourgoin, 2010) may exacerbate risks for PWSUDs and PWIDs (Vasylyeva et al.; Ying, Yang, & Jianming, 2020). Drug supply chains may be disrupted, and changes in licit and illicit markets may be accompanied by reductions in quality and safety (Knopf, 2020; Nagelhout et al., 2017; Rowe et al., 2016; EMCDDA, EUROPOL, 2020).

Furthermore, patients’ accessibility to treatment services could be restricted due to lockdown policies (Bojdani et al., 2020; Roncero et al, 2020). Patients receiving opioid agonist treatment (OAT) may not be able to access daily doses of medications (Arya & Gupta, 2020); spatial distancing may make home detoxification difficult; closing of non-essential services and utilising staff and other resources to manage acute COVID-19 cases could result in sudden and uncoordinated closures of services for PWSUDs (EMCDDA, 2020). Individuals who use multiple substance may be particularly impacted (Mellis AM, In press). Adaptive capacities of systems to epidemic situations that need coordinated responses may relate directly to vulnerabilities of the same systems (Smit & Wandel, 2006). Accessibility to and equal distribution of wealth (financial and other resources, reliable and correct information and communication channels, appropriate and proportionate working technologies) compounded by reductions in social and relationship capital may impact social resilience to coping with pandemics (Dolan & Walker, 2006).

To understand better complexities that are challenging addiction treatment and harm reduction services globally, the International Society of Addiction Medicine (ISAM) has been conducting a longitudinal global survey aiming to evaluate rapidly and over time how different countries are maintaining and/or reorganizing their substance use treatment and harm reduction services during the COVID-19 pandemic. This paper will report on how different countries have adapted their health system response to emerging needs in the first month after the official announcement of the pandemic by the WHO.

## 2. Methods

Description of the methodology used for this survey has been published as a study protocol (Baldacchino et al., 2020). Potential respondents were contacted on April 4^th^, 2020 asking about the COVID-19 pandemic impact on PWSUDs in their own countries. Data collection was concluded on May 8^th^, 2020.

### 2.1. Questionnaire

The questionnaire consisted of 92 questions in two main areas: (1) situation assessment during the pandemic; and, (2) health responses to the pandemic. This paper will focus on health responses during the COVID-19 pandemic period (Baldacchino et al., 2020). Results on the situation assessment is reported in another publication (Farhoudian, Radfar, et al., 2020).

Questions around health responses to the pandemic were grouped into 3 categories:

1. Systems available to respond to acute emerging needs due to the COVID-19 pandemic within substance use services;
2. Availability of protocol and/or guidelines around COVID-19 and PWSUDs; and,
3. Reduction in face-to-face contacts as a result of lockdown policies.

To assess respondents’ overall views, they were asked to score the “overall situation at a glance” rating scale questions (RSQ) (between 1 to 10 with 1 for the worst situation and 10 for the best situation) based on their opinion regarding the overall quality of the situation of their country for each of the above 3 sections.

### 2.2. Categorization of countries based on their income

The 2019 statistical annex of World Economic Situation and Prospects (WESP) (“World Economic Situation and Prospects 2019,” 2019) was used to categorize responding countries. Very low- and low-income categories were merged into one, retaining middle- and upper-income countries designations. In figures, countries’ names are sorted alphabetically in each group of high-, middle- and low-income categories. The number of respondents (for countries with more than one respondent) is indicated in front of their names, and numbers in each column represent valid responses from each country.

### 2.3. Statistical Analysis

Statistical analyses were performed using SPSS version 22 (IBM Corp., Armonk, N.Y., USA) and RStudio (version 1.2.1335). Descriptive data are presented as means and percentages for each country’s response mean (percentage), as well as an average to the global responses.

### 2.4. Ethics Approval

The survey protocols and all materials, including the survey questionnaires, received approval from the University of Social Welfare and Rehabilitation Sciences, ethics committee in Tehran, Iran (Code: IR.USWR.REC.1399.061).

## 3. Results

### 3.1. Participants

A total of 177 respondents from 77 countries participated. Figure 1 shows the distribution of the countries and number of participants from each. Among 177 respondents, 95 (53.7%) were from high-income, 34 (19.2%) from middle-income, and 48 (27.1%) from low-income countries (“World Economic Situation and Prospects 2019,” 2019). Table 1 shows respondents’ demographic characteristics classified by their associated countries’ income.

**Table 1.**
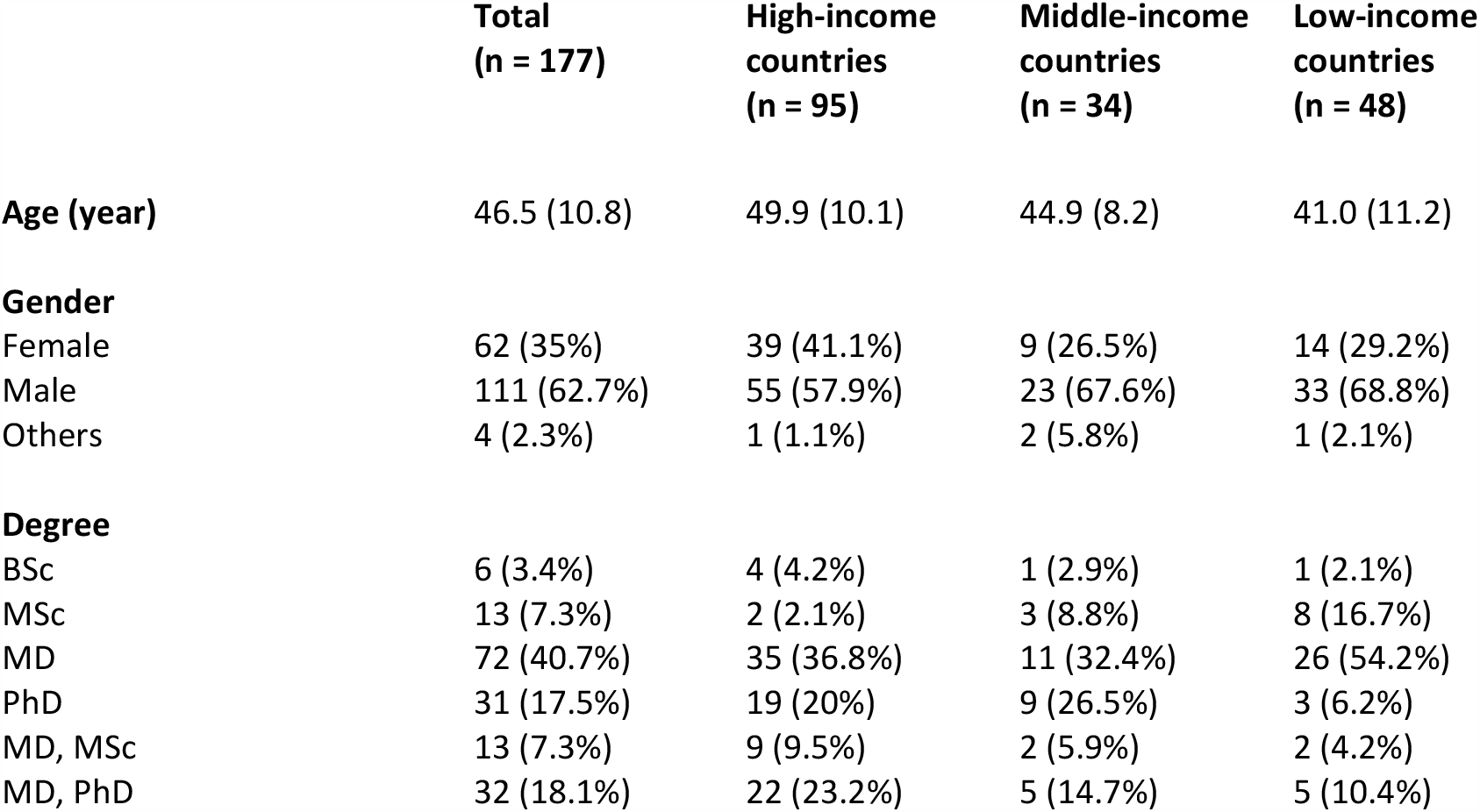
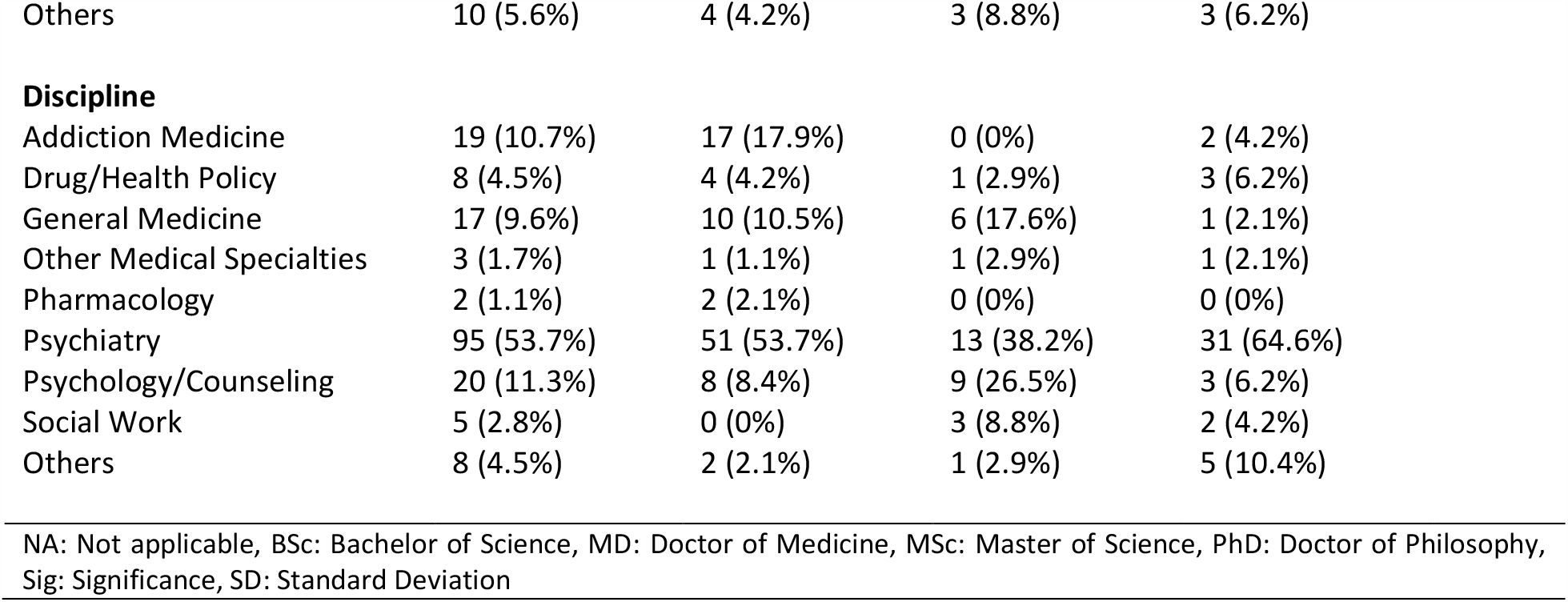
Survey respondents’ demographic, educational, and professional information classified by their countries’ income status. Variables are reported as mean (standard deviation) or count (percent%).

|  | **Total**<br>**(n = 177)** | **High-income**<br>**countries**<br>**(n = 95)** | **Middle-income**<br>**countries**<br>**(n = 34)** | **Low-income**<br>**countries**<br>**(n = 48)** |
| --- | --- | --- | --- | --- |
| **Age (year)** | 46.5 (10.8) | 49.9 (10.1) | 44.9 (8.2) | 41.0 (11.2) |
| **Gender** |  |  |  |  |
| Female | 62 (35%) | 39 (41.1%) | 9 (26.5%) | 14 (29.2%) |
| Male | 111 (62.7%) | 55 (57.9%) | 23 (67.6%) | 33 (68.8%) |
| Others | 4 (2.3%) | 1 (1.1%) | 2 (5.8%) | 1 (2.1%) |
| **Degree** |  |  |  |  |
| BSc | 6 (3.4%) | 4 (4.2%) | 1 (2.9%) | 1 (2.1%) |
| MSc | 13 (7.3%) | 2 (2.1%) | 3 (8.8%) | 8 (16.7%) |
| MD | 72 (40.7%) | 35 (36.8%) | 11 (32.4%) | 26 (54.2%) |
| PhD | 31 (17.5%) | 19 (20%) | 9 (26.5%) | 3 (6.2%) |
| MD, MSc | 13 (7.3%) | 9 (9.5%) | 2 (5.9%) | 2 (4.2%) |
| MD, PhD | 32 (18.1%) | 22 (23.2%) | 5 (14.7%) | 5 (10.4%) |
| Others | 10 (5.6%) | 4 (4.2%) | 3 (8.8%) | 3 (6.2%) |
| Discipline |  |  |  |  |
| Addiction Medicine | 19 (10.7%) | 17 (17.9%) | 0 (0%) | 2 (4.2%) |
| Drug/Health Policy | 8 (4.5%) | 4 (4.2%) | 1 (2.9%) | 3 (6.2%) |
| General Medicine | 17 (9.6%) | 10 (10.5%) | 6 (17.6%) | 1 (2.1%) |
| Other Medical Specialties | 3 (1.7%) | 1 (1.1%) | 1 (2.9%) | 1 (2.1%) |
| Pharmacology | 2 (1.1%) | 2 (2.1%) | 0 (0%) | 0 (0%) |
| Psychiatry | 95 (53.7%) | 51 (53.7%) | 13 (38.2%) | 31 (64.6%) |
| Psychology/Counseling | 20 (11.3%) | 8 (8.4%) | 9 (26.5%) | 3 (6.2%) |
| Social Work | 5 (2.8%) | 0 (0%) | 3 (8.8%) | 2 (4.2%) |
| Others | 8 (4.5%) | 2 (2.1%) | 1 (2.9%) | 5 (10.4%) |

NA: Not applicable, BSc: Bachelor of Science, MD: Doctor of Medicine, MSc: Master of Science, PhD: Doctor of Philosophy, Sig: Significance, SD: Standard Deviation

**Figure 1.**
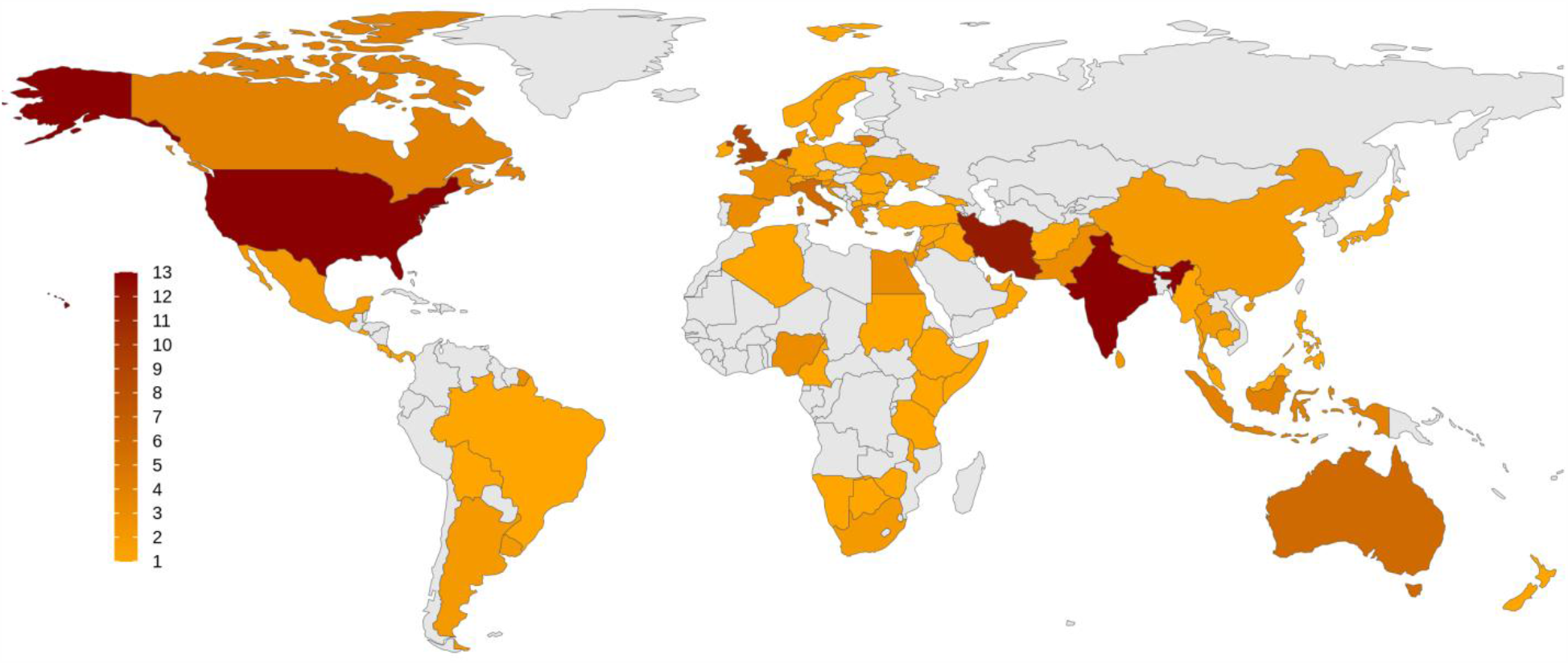
Global distribution of the respondents to the survey. Number of participants from each country is demonstrated as a color spectrum from light to dark purple.

### 3.2. Implementing business continuity/contingency plans

Among respondents from high-income countries (N=95), 69% answered that business continuity/contingency plans had been implemented in their countries to make sure that services continued to operate for PWSUDs during the COVID-19 pandemic compared to 40.7% in middle-income (N=34) and 53.8% (N=48) in low-income countries. Overall, respondents from 56% of participating countries reported that business contingency plans had been arranged to help ensure that continuity of services during the pandemic (Figure 2).

**Figure 2.**
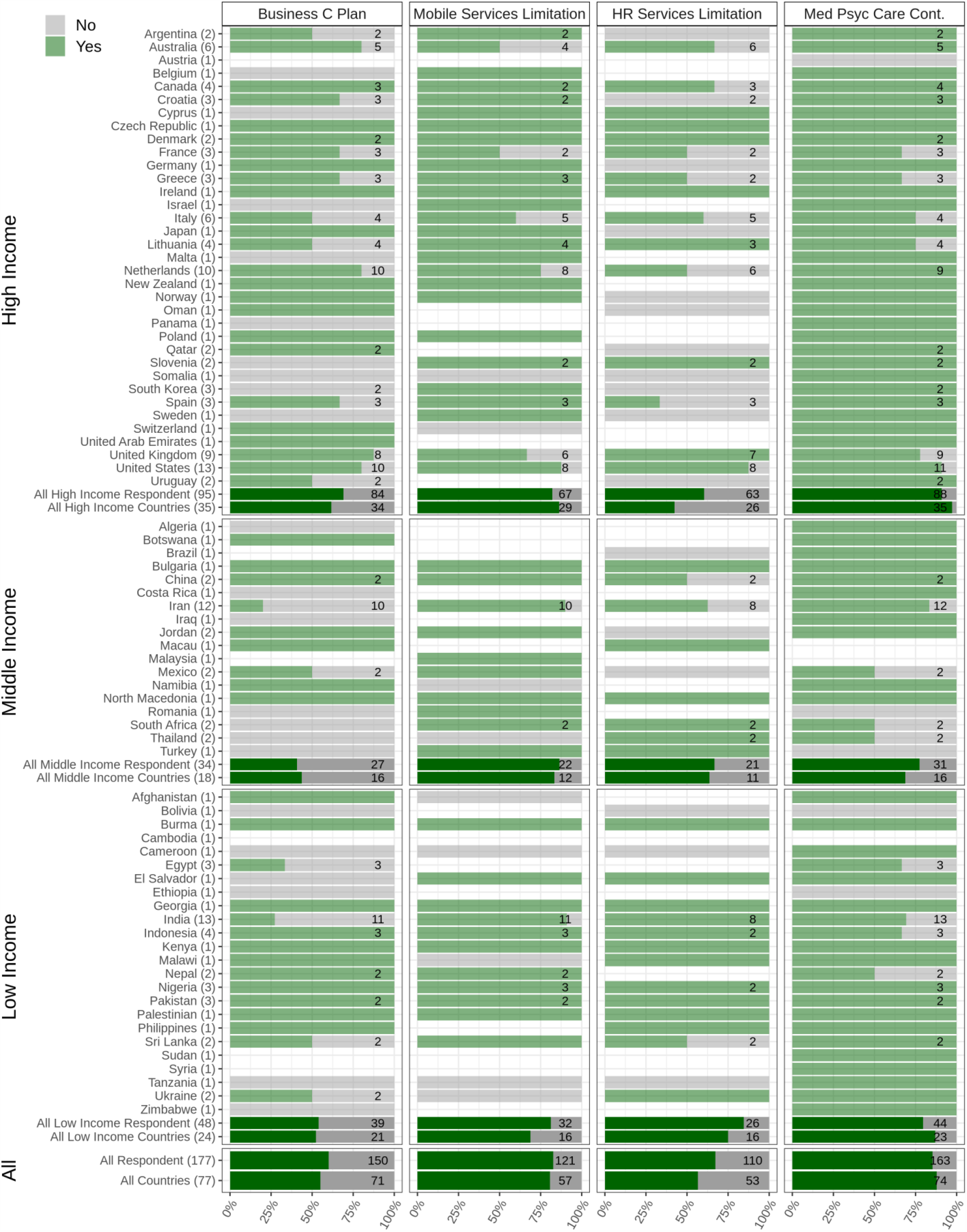
Availability and accessibility of treatment and harm reduction services. Data relating to arranging business continuity plans (Business C Plan), limitations that mobile services faced during the pandemic (Mobile services limitation), limitations that harm reduction services faced during the pandemic (HR services limitation), and continuity of other medical and psychiatric necessary care (Med Psyc Care Cont.) are depicted. The figure shows responses from from respondents from 77 countries categorized into low-income, middle-income and high-income countries. The light green bars and the numbers associated with each country show the survey respondents who reported having experienced limitations regarding the question in their country (Yes), and the grey bars shows the survey respondents who reported having experienced no limitations regarding the question in their country (No). The dark green bars show the overall responses in each category (low-income, middle-income and high-income) as well as overall responses.

### 3.3. Availability and accessibility of treatment and harm reduction services

Among respondents from high-income countries (N=95), 57% answered that treatment and harm reduction services for PWSUDs had been available and accessible in their countries during the pandemic onset compared to 51.6% in middle-(N=34) and 63% in low-(N=48) income countries. Overall, respondents from 59% of participating countries reported that treatment and harm reduction services for PWSUDs had been available and accessible during the initial period of the COVID-19 pandemic (Figure 2).

Respondents from over 81% of participating countries (N=77) reported having experienced limitations in the usage of any outreach services due to lockdown policies for homeless PWSUDs. Furthermore, respondents from 57% of participating countries reported having experienced limitations in their harm reduction overdose services during the initial period of the pandemic. Problems with the distribution of take-home naloxone were reported by respondents from 57% of participating countries. Respondents from 54.8% of the participating countries reported shortages in needle and syringe programs (NSPs) and/or with respect to condom distribution.

### 3.4. Medical and psychiatric care during the initial period of the pandemic

Among respondents from high-income countries (N=95), 90% answered that medical and psychiatric care for PWSUDs had been available during the initial stages of the pandemic compared to 77.4% in middle-income (N=34) and 79.5% in low-income (N=48) countries. Overall, respondents in 88% of participating countries reported that necessary medical and psychiatric care for PWSUDs had continued in their countries during this period (Figure 2). However, respondents in 37.5% of participating countries reported having experienced shortages of opioid medications (methadone or buprenorphine).

Only 44.3% of respondents from high-income (N=95), 32.2% from middle-income (N=34) and 40.1% from low-income (N=48) countries reported that COVID-19 screening and/or diagnosis test kits based on local/national guidelines for PWSUD had been available in their country. Overall, respondents from only 48% of the participating countries reported that there had been enough personal protective equipment (PPE) available for PWSUDs during the initial stage of the pandemic. Respondents from 77.7% of participating countries reported SUD health workers’ safety as a concern for employers in the outpatient treatment centers, 66.4% had received training regarding their safety and 72.9% reported that they had had access to enough PPE.

Distribution of other responses on the effect of COVID-19 on substance use treatment and/or harm reduction services to vulnerable groups such as children, women, pregnant women and immigrants or refugees can be seen in Table 2 and Figure 4. Table 2 shows existence of services for children, women, pregnant women and refugees or immigrants among the countries based on their income group.

**Table 2.** Services for children, women, pregnant women and refugees or immigrants among the countries based on their income group. Availability of the services is reported in part a. Continuity of the service as usual or with limitations among countries which have the service available is reported in b and c parts.

| Target group | Total<br>% (n) | High income<br>countries<br>% (n) | Middle income<br>countries<br>% (n) | Low income<br>countries<br>% (n) |
| --- | --- | --- | --- | --- |
| **a. Service Availability** |  |  |  |  |
| Children | 80.8 (130) | 79.4 (63) | 92.3 (26) | 75.6 (41) |
| Women | 95.4 (153) | 96.3 (81) | 96.6 (30) | 92.8 (42) |
| Pregnant Women | 88 (149) | 88.4 (78) | 89.3 (28) | 86 (43) |
| Immigrants/<br>Refugees | 70.1 (124) | 68.2 (63) | 82.6 (28) | 65.8 (34) |
| **b. Continued as Usual** |  |  |  |  |
| Children | 22.8 (30) | 18 (12) | 16.6 (5) | 35.5 (15) |
| Women | 21 (33) | 16.6 (14) | 20.7 (6) | 28.2 (12) |
| Pregnant Women | 28.2 (42) | 23.2 (18) | 28 (8) | 37.8 (16) |
| Immigrants/<br>Refugees | 18.4 (23) | 11.6 (8) | 21 (6) | 28 (10) |
| **c. Continued with Limitations** |  |  |  |  |
| Children | 77.2 (100) | 82 (51) | 83.3 (21) | 64.5 (26) |
| Women | 79 (120) | 77.2 (67) | 83.4 (24) | 79.3 (30) |
| Pregnant Women | 71.8 (107) | 76.8 (60) | 72 (20) | 62.2 (27) |
| Immigrants/<br>Refugees | 81.6 (101) | 88.4 (55) | 79 (22) | 72 (24) |
| **% has been calculated based on Yes response in the respondents in each group of income** |  |  |  |  |

**Figure 3.**
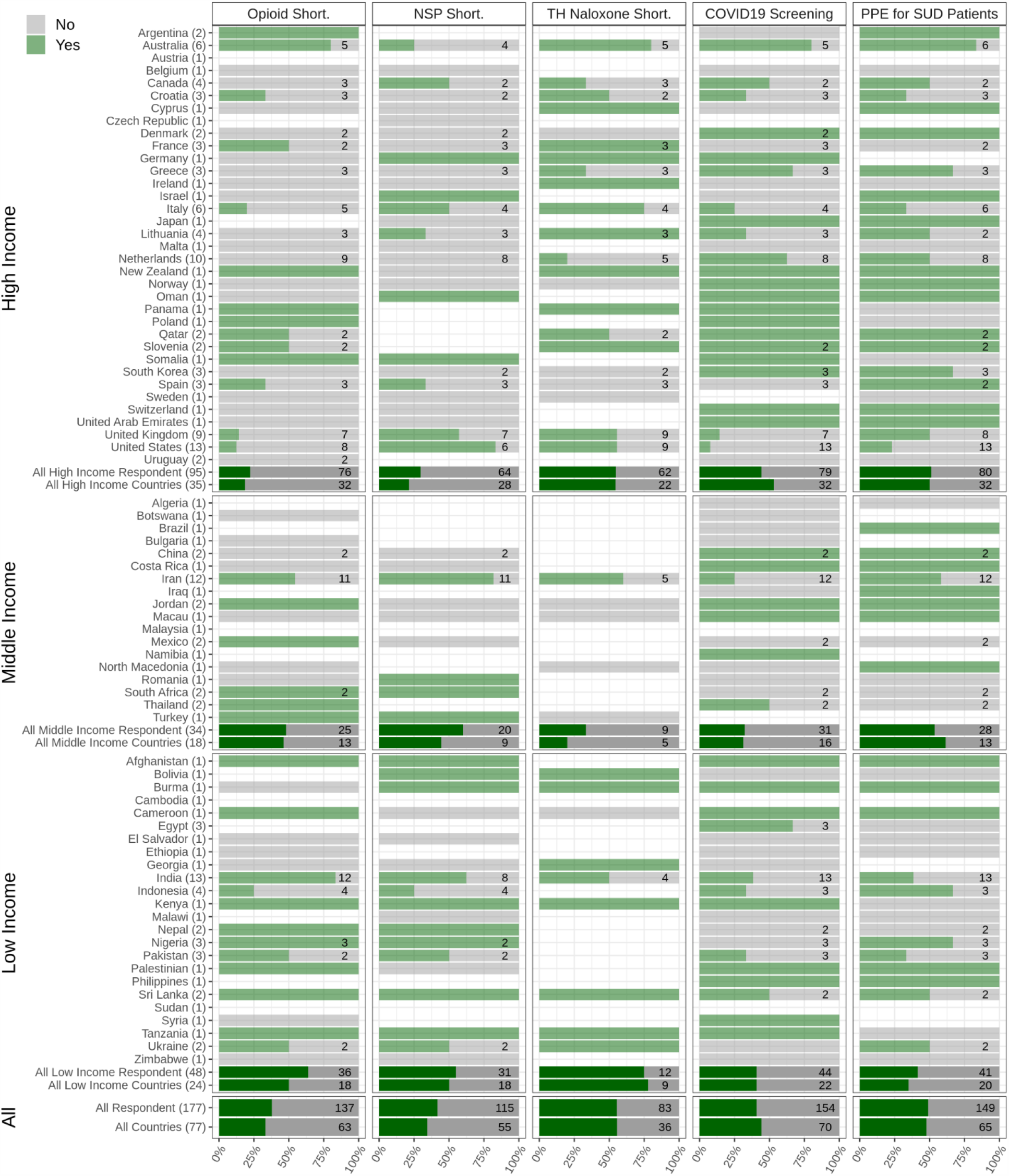
Medical services for people with substance use disorders during the pandemic. The responses of respondents from 77 countries are shown, categorized into low-income, middle-income and high-income countries to the questions related to the shortages in opioid medication (opioid short.), disruption in needle and syringe and/or condom distribution services (NSP Short.), availability or shortages in take-home naloxone services (TH Naloxone short.), availability of COVID-19 screening kits and equipment for PWSUDs in their countries (COVID19 screening), and personal protective equipment provision to PWSUDs (PPE for SUD patients).

**Figure 4.**
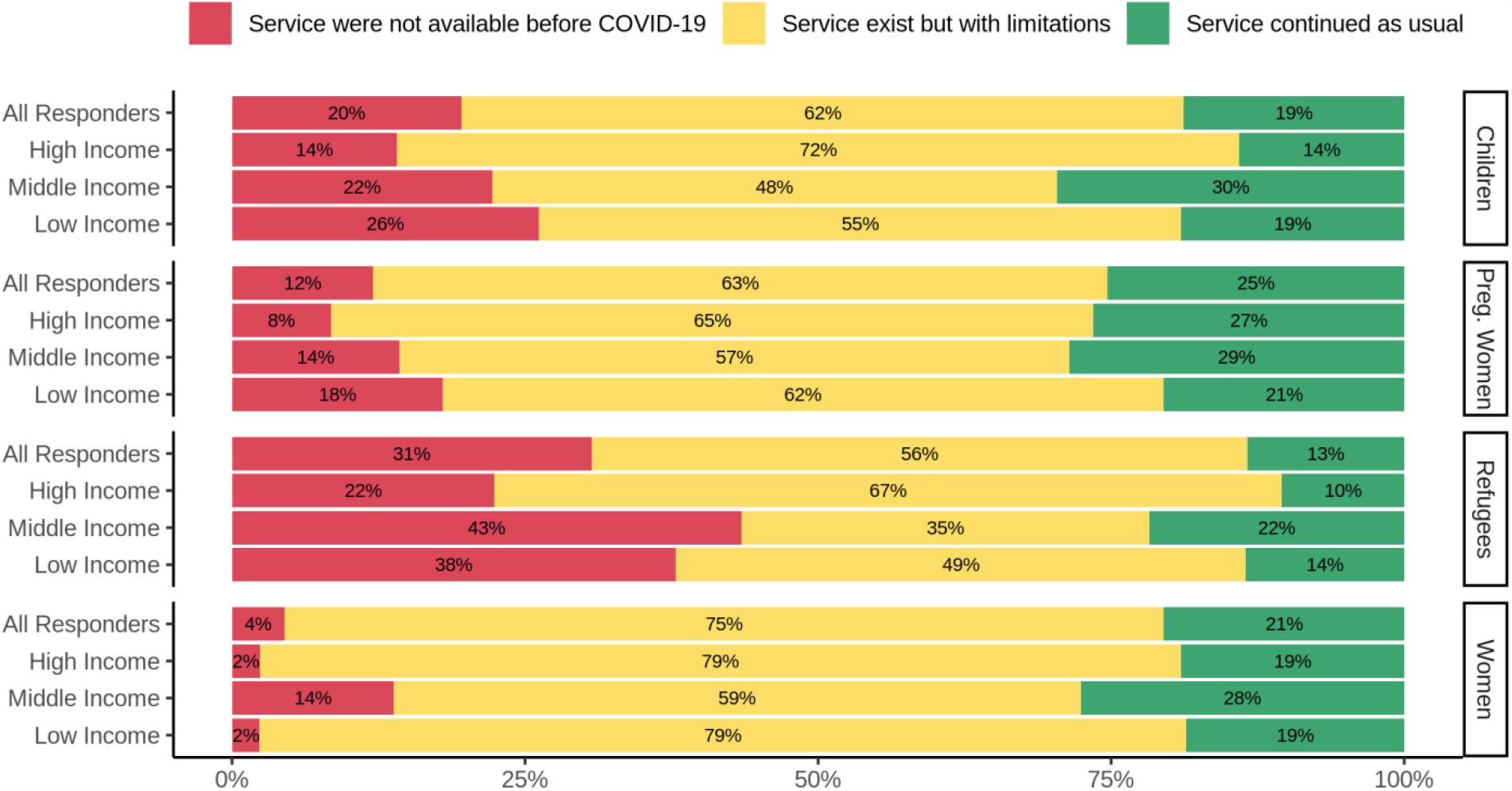
Effects of COVID-19 on Substance Use Treatment and/or Harm Reduction Services for Vulnerable Groups. Services for children, pregnant women, refugees, and women, in high, middle- and low-income countries are depicted. The red, yellow, and green bars depict the responses indicating lack of availability of services during the COVID-19 pandemic, existence of limited services, and usual service provision, respectively.

Overall, 22.8% of all respondents replied that service for children continued as usual compared to 77.2 % that replied service for children continued but with limitations. According to the responses, in all three groups of income countries treatment and/or harm reduction services for pregnant women were a group with minimum impact from COVID-19. Refugees and immigrant population was the group that their services impacted more than other groups due to COVID-19. Only 18.4% replied that service for refugees and/or immigrants population continued as usual and 81.6% replied that this service continued but with limitations.

### 3.5. Health Policies for COVID-19 among PWSUDs

Overall, respondents from 48% of the participating countries reported the presence of local and/or national guidelines tailored to be used during the initial stage of the pandemic (60.2% in high-income, 57.1% in middle-income and 29% in low-income countries). Among respondents from high-income countries, 65.7% answered that there had been a protocol available for COVID-19 screening in different sectors of treatment for PWSUDs or harm reduction facilities compared to 60% in middle-income and 82.3% in low-income countries.

Over 76% of respondents from high-income, 63.3% from middle-income and 63% from low-income countries reported that there had been guidelines available that helped service providers in the management and/or referral of PWSUDs with symptoms of COVID-19.

Most respondents replied that there had been plans to restrict personal contacts and decrease patients’ commutes for treatment in their countries (86%, 90%, 86.6% in high-, middle- and low-income countries, respectively and 85% overall) due to their national and regional lockdown policies.

As a result, respondents from 80% of the participating countries reported that clinicians had been prescribing longer-period prescriptions (e.g., 28 days rather than weekly) to PWSUDs during the onset of the pandemic (Figure 5).

**Figure 5.**
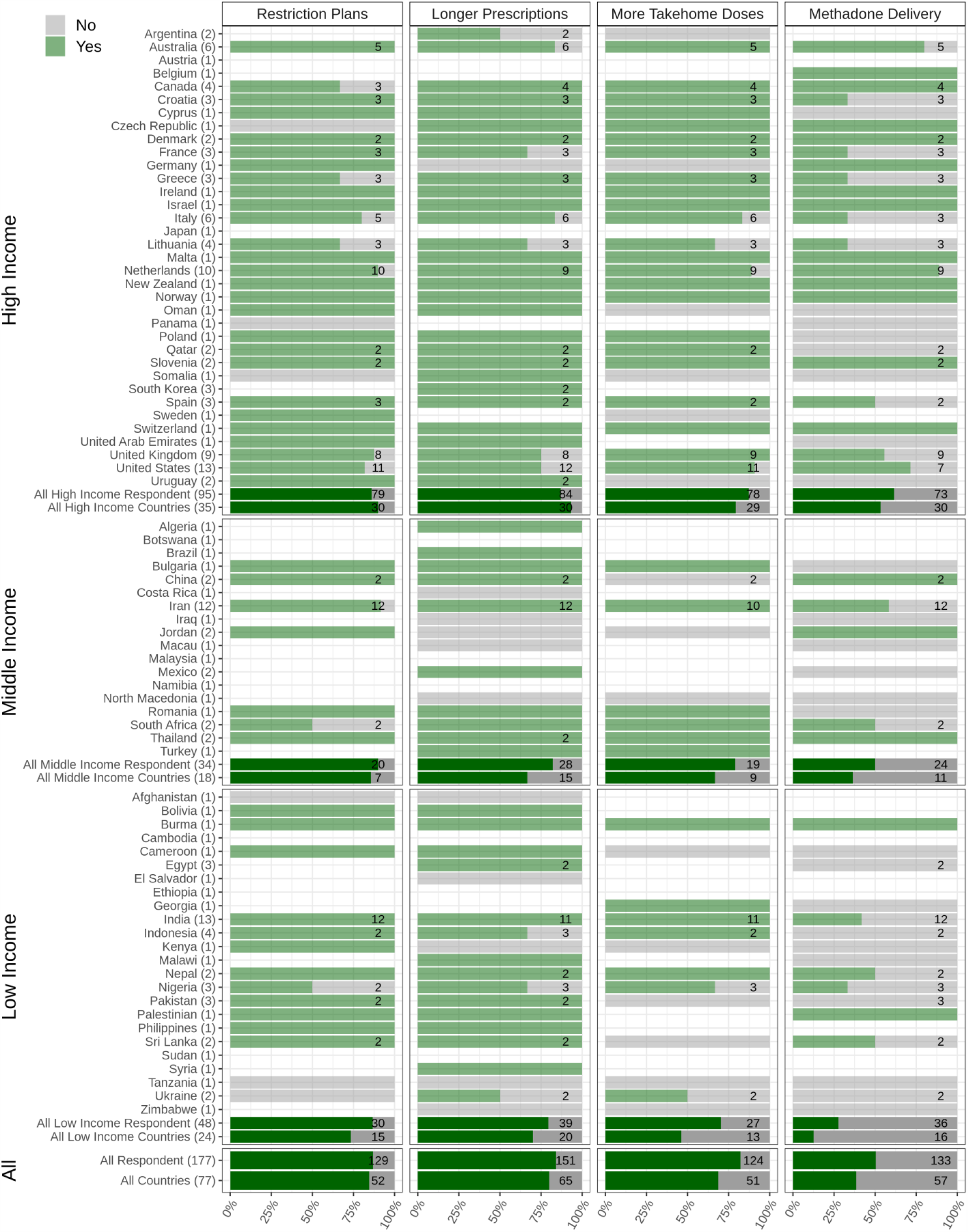
Health Policies for COVID-19 among PWSUDs. Plans to restrict any personal contact, provision of prescriptions of longer durations, provision of more take-home doses of opioids drugs and availability of any program for delivering opioid drugs to patients’ homes are depicted. The figure shows responses from 77 countries which are categorized into low-income, middle-income and high-income. The light green bars and the numbers associated with each country shows the survey respondents who reported having experienced limitations regarding the question in their country (Yes), and the grey bars shows the survey respondents who reported having experienced no limitations regarding the question in their country (No). The dark green bars show the overall responses in each category (low-income, middle-income and high-income) as well as overall responses.

Additionally, around 69% of participating countries reported that clinicians within OAT programs had provided more take-home doses of methadone and/or buprenorphine during the onset of the pandemic. Regionally, 61.6% of respondents from high-income, 50% from middle-income and 27.7% from low-income countries reported this approach had been used in their countries (Figure 5).

Respondents from high-income countries most frequently reported having had availability of long-acting injectable buprenorphine (34.9%; n=63). Overall, respondents from 22% of participating countries reported that long-acting injectable Buprenorphine had been available as a therapeutic option.

Figure 6 shows the average score of each question based on income categorization. Maximum contrast between high- and low-income countries was seen in the availability and access to treatment and harm reduction services. Maximum and minimum differences between high- and middle-income countries were observed in flexibility in service provision and countries’ reactions to the COVID-19 pandemic, respectively.

**Figure 6:.**
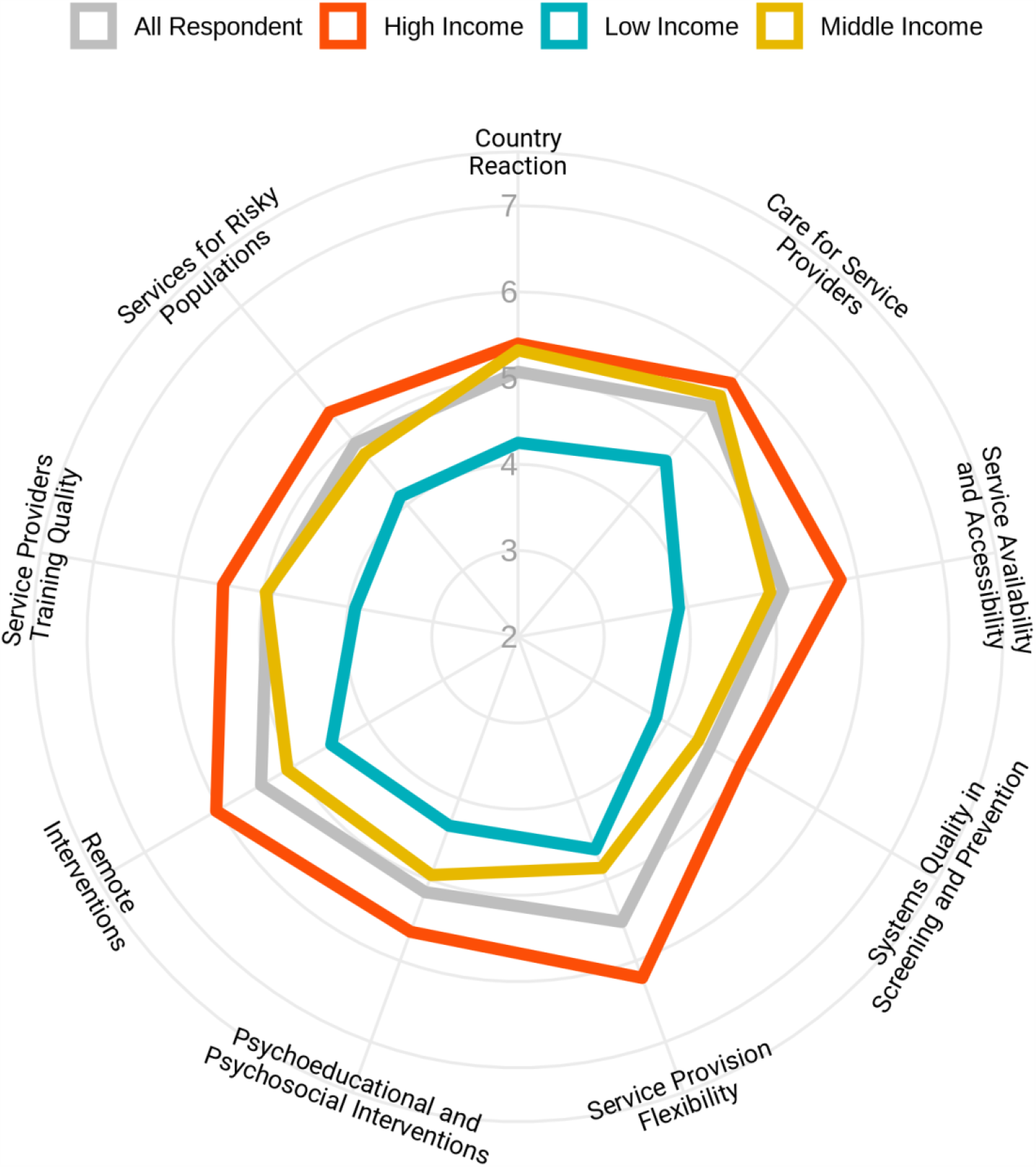
Flexibility of health responses for people with substance use disorders in response to the pandemic in different domains based on the income levels of the countries. Respondents were asked to rate the overall flexibility of their health system in 9 different domains from 1 (extremely poor) to 10 (extremely good).

An average for all rating scale questions in different domains has been calculated, and Figure 7 shows the results in a global map format.

**Figure 7.**
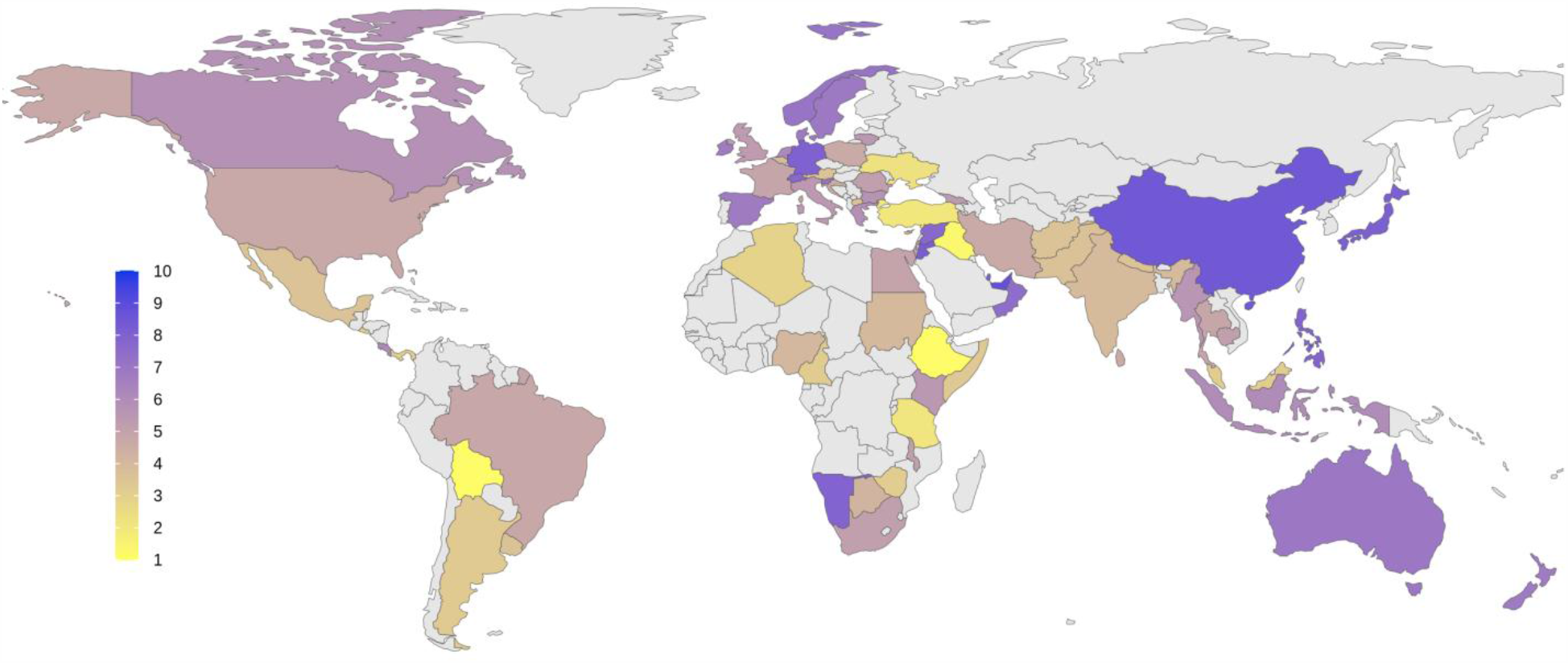
Overall quality of health response to COVID-19 pandemic based on the subjective ratings by respondents from different countries. Average scores were measured based on responses in 9 domains depicted in Figure 6. Score 1 represents for the worst quality in response and 10 represents the best situation in favor of health services. Average scores for each country are shown using a color spectrum from yellow to blue.

## 4. Discussion

The emergence of COVID-19 in early 2020 raised considerable challenges for substance use treatment and harm reduction programs around the world as reflected in this global survey. The need for effective spatial distancing and isolation to protect patients, the treatment workforce, and people in contact with patients and health workers has placed increased demands on treatment services provision, with potential imbalances in impact on particularly vulnerable patient populations (Mellis et al., in press). Here, in this global survey we have explored different challenges and health responses in 77 countries. Our findings showed that respondents from 56% of participating countries reported business contingency plans had been arranged to help ensure that services would continue to operate during the pandemic, which is compatible with responses to another question indicating that 41% of respondents believed there had not been sufficient availability and accessibility of treatment and harm reduction services during the onset of the pandemic in their countries at the time of survey completion.

As a preventative measure to reduce COVID-19 spread, all international and national published guidelines advised limited but effective ways regarding how to initiate treatment, support stabilization and maintenance and continue to provide harm reduction measures to treatment-seeking and other populations with substance use problems (Farhoudian, et al., 2020; Marsden, et al., 2020). These recommendations often included extending flexibility in OAT services with reduced supervision of doses and increased home delivery (Dunlop et al., 2020; Peavy et al., 2020). Another step taken to adjust to the present situation included expanding telemedicine and teletherapy services (Farhoudian, Baldacchino, et al., 2020; Knopf, 2020; Elizabeth A Samuels et al., 2020; Mellis et al., in press).

The COVID-19 pandemic issynergistic interacting with a substance use epidemic globally creating a *syndemic* (defined as a synergistic epidemic, the aggregation of two or more concurrent or sequential epidemics, which exacerbate the prognosis and burden of disease (Singer, Bulled, Ostrach, & Mendenhall, 2017)). During the COVID-19 pandemic, marginalized people including PWSUDs are at greater risk of increased morbidity and mortality (Dorahy et al., 2015). These syndemically disadvantaged populations may be more likely to experience disparate, possibly substandard, service provision in systems prioritizing resource needs around a pandemic response (Inverse Response Law and Inverse Care Law) (Phibbs, Kenney, Rivera-Munoz, & Huggins, 2018). Such inequities may present at macro levels around effective and appropriate policymaking at national, organizational, and local levels (Phibbs et al., 2018; Watt, 2002) and at micro levels around areas of access to resources, social services, public health benefits of medical treatments, pharmacies, health care facilities and provision of medical equipment (Davis, Wilson, Brock-Martin, Glover, & Svendsen, 2010; Runkle, Brock-Martin, Karmaus, & Svendsen, 2012; Watt, 2002).

Proactive business continuity plans for PWSUDs are important for all governments as part of COVID-19 re-mobilization plans and possible future responses to similar pandemics to support and avert delays and inequities in responses. PWSUDs are at risk for negative impact of COVID-19 (Khatri & Perrone, 2020; Volkow, 2020), and our findings showed that 88% of respondents reported continuity of other necessary medical and psychiatric care compared to less than 60% who reported existence of business continuity/contingency plans and enough availability and accessibility of treatment and harm reduction services for PWSUDs. These findings suggest that policymakers and health authorities in each country could have possibly made more appropriate decisions in order to protect at-risk and marginalized PWSUDs including those who may be homeless, have HIV/AIDS, hepatitis, or multiple and complex morbidities. Such decisions may involve considering how best to provide scheduled and new appointments and prescription medications in the circumstances of lockdowns.

Based on our finding in this global survey, we provide the following recommendations.

### RECOMMENDATION 1

International organizations such as the WHO and United Nations Office of Drug Control (UNODC) and other related groups such as the International Society of Addiction Medicine (ISAM), International Society of Substance Use Professionals (ISSUP), and World Federation Against Drugs (WFAD) should provide adequate support to raise policymakers’ knowledge in the area of addiction medicine on how to establish business continuity committees during initial stages of pandemics in order to make advanced care planning decisions through effective leadership.

Continuity of services especially in crisis situations needs certain, evidence-based, and locally tailored protocols and guidelines. In our study, addiction medicine professionals reported that most of their countries did not provide early guidelines or protocols to tailor their services to the pandemic. It is important to consider that respondents in only one-third of low-income countries reported the availability of such guidelines compared to respondents in half of high-income countries. Another survey (Mongan, Galvin, Farragher, Dunne, & Nelson, 2020) conducted in four high income regions (New South Wales, Ireland, Scotland, New York State, and British Columbia) found that special guidelines in response to the new situation and assurance of continuity of the services were available very soon after the start of lockdown, which is consistent with our findings that high income countries had a more timely response in this domain. In the absence of guidelines and protocols, clinicians and service providers may not effectively balance different competing ethical and professional issues when they are making clinical and operational decisions when many things may be happening that could potentially be conflicting in nature (e.g. maintaining stability but reducing therapeutic contacts). Guidelines also allow stakeholders to improvise and identify innovative ways through evidence-based solutions to help decrease the dual burden of substance use and COVID-19 infection (Samuels et al., 2020; Sokol, Gupta, Powers, Hoffman, & Meza, 2020).

### RECOMMENDATION 2

Governments and local authorities should be cognizant that an effective response system is based around a well informed and supportive environment. Available and communicated international and national clinical guidelines are pivotal in future responses to similar pandemics when supporting PWSUDs.

The World Drug Report 2020 stated that, “*If Governments respond the same way to the current economic slump, interventions such as prevention of drug use and related risk behaviors and drug treatment services could be hard hit*” (Nations, 2020). Substance use accounts for approximately 11% of the global health burden (Forouzanfar et al., 2016). Treatment is one important strategy for reducing the burden of disease. A study of World Mental Health Surveys (Organization, 2010) found that only 7.1% of PWSUDs had received at least minimally adequate treatment in the past year (10.3%, 4.3% and 1.0%, respectively, in high-, upper-middle, and low/lower-middle income countries)(Degenhardt et al., 2017). Poor access to treatment, awareness/perceived treatment need, and compliance (on the part of both provider and client) have been reported to be main barriers for substance use treatment (Degenhardt et al., 2017).

Our results showed that shortage of opioid medication for maintenance treatment was reported by respondents from about 40% of participating countries. Lack of opioid medications in patients undergoing maintenance treatment is a risk factor for lapse, relapse and/or overdoses. This situation may become more severe when transport and other supply chains are disrupted compounded with reduced provision by pharmacies and other dispensing outlets either due to spatial distancing, reduced hours of service and/or closing during the pandemic.

### RECOMMENDATION 3

International organizations with regional and local government structures should create contingencies around adequate supplies of medications such as methadone and buprenorphine. Harm reduction services, especially outreach services, are among the most effective strategies for prevention of HIV, HCV and HBV transmission among the most at risk populations (Nazari et al., 2016; Needle et al., 2005; Peak, Rana, Maharjan, Jolley, & Crofts, 1995). Harm reduction services seem to be among the most affected during the initial stages of the COVID-19 pandemic. Eighty-one (81) % of participating countries reported limitations in usage of any mobile and other outreach services due to lockdown policies for homeless PWSUDs, with respondents from 57% of participating countries reporting limitations in their harm reduction overdose services during the initial period of the pandemic. This was compounded with reported problems with the distribution of take-home naloxone as reported by respondents from 57% of participating countries. Finally, respondents from 54.8% of participating countries reported that there have been shortages at needle and syringe programs and/or of condom distribution.

### RECOMMENDATION 4

Harm reduction initiatives should be seen as an integral part of an evidence-based treatment program and not as an adjunct to failed treatment and/or solely as a public health response to reduce blood-borne diseases. Service providers should be considering identifying person-centered, continuous care provision in all therapeutic options available (harm reduction initiatives included) especially during pandemic situations

Pregnant women and immigrants /refugees with SUDs are particularly vulnerable groups. According to survey responses, pregnant women were perceived as relatively less impacted during the initial period of the pandemic. This is reassuring as discontinuity of treatment services could place not only a pregnant woman at high risk, but also the developing fetus. However, refugee and immigrant populations were reported as having had their services impacted more than other groups due to the pandemic. Only 12.9% of respondents replied that service for refugees and/or immigrants population continued as usual, and 57.3% replied that this service continued but with severe limitations.

### RECOMMENDATION 5

Vulnerable groups such as immigrants and refugees with SUDs should have access to all possible therapeutic options available as described in the UN charter in the Human Right Convention (“International Convention on the Protection of the Rights of All Migrant Workers and Members of Their Families”). It is important that appropriate evidence-based services are designed and implemented by health authorities for such vulnerable groups.

Availability of all relevant resources is essential in the delivery of quality services. Our findings suggest that in general in multiple domains of countries’ reactions to the pandemic (e.g., availability of and access to treatment and harm reduction, screening and early interventions, flexibility in service provision and services for special and high-risk populations), the COVID-19 pandemic has had more negative impact that is linked to the income level of countries.

This study has multiple limitations. The responses obtained was intentionally based around personal opinions of addiction medicine experts to help understand the “state of things in real life” rather than objective epidemiological data which would have been considerably delayed. The limited number of respondents makes this information non-representative and possibly biased. Given the urgency of the COVID-19 pandemic, the aim of the paper is to alert and inform colleagues around the world and facilitate collaboration. Due to the time limitations, the questionnaire was circulated only in English. Therefore, some experts may have withdrawn from the survey for lingual reasons and others may have answered questions less precisely.

Addiction medicine systems in all countries, regardless of income level, have been affected to some degree by the COVID-19 pandemic. Depending on the different domains and the ability of countries to adapt to existing conditions, these effects may differ across jurisdictions. Income level may relate importantly to responses and impact vulnerable groups like PWSUDs. Our recommendations will hopefully support a more resilient system of care that improves responses to future COVID-19 waves and other pandemics.

## Data Availability

The full description of the research design and analysis plan is available upon request.

## Declarations

Authors declare no conflict of interests. Anja Busse is a staff member of UNODC. The authors alone are responsible for the views expressed in this article and they do not necessarily represent the decisions or policies of the UNODC.

## Acknowledgment

Authors would like to thank Arash Khojasteh Zonoozi and Hossein Mohadess Ardebili for their insightful comments into the initial draft of the manuscript.

## References

Adger, W. N. (1999). Social vulnerability to climate change and extremes in coastal Vietnam. World development, 27(2), 249–269.

Ahern, J., Stuber, J., & Galea, S. (2007). Stigma, discrimination and the health of illicit drug users. Drug and alcohol dependence, 88(2-3), 188–196.

Arya, S., & Gupta, R. (2020). COVID-19 outbreak: Challenges for Addiction services in India. Asian Journal of Psychiatry, 51, 102086.

Baldacchino, A., Radfar, S. R., De Jong, C., Rafei, P., Yunesian, M., Gerra, G., … Khojasteh Zonoozi, A. (2020). COVID-19 and Substance Use Disorder: Study Protocol for the International Society of Addiction Medicine Practice and Policy Interest Group Global Survey. Basic and Clinical Neuroscience, 11(2), 155–162.

Bhaskaram, P. (2002). Micronutrient malnutrition, infection, and immunity: an overview. Nutrition reviews, 60(suppl_5), S40–S45.

Bojdani, E., Rajagopalan, A., Chen, A., Gearin, P., Olcott, W., Shankar, V., … Batty, N. (2020). COVID-19 Pandemic: Impact on Psychiatric Care in the United States, a Review. Psychiatry Research, 113069.

Cheng, S. K., & Lam, D. J. (1997). Relationships among life stress, problem solving, self-esteem, and dysphoria in Hong Kong adolescents: Test of a model. Journal of Social and Clinical Psychology, 16(3), 343–355.

Clay, J. M., & Parker, M. O. (2020). Alcohol use and misuse during the COVID-19 pandemic: a potential public health crisis? The Lancet Public Health, 5(5), e259.

Coetzee, B. J. s., & Kagee, A. (2020). Structural barriers to adhering to health behaviours in the context of the COVID-19 crisis: Considerations for low-and middle-income countries. Global Public Health, 1–10.

Cook, R. T. (1998). Alcohol abuse, alcoholism, and damage to the immune system—a review. Alcoholism: Clinical and Experimental Research, 22(9), 1927–1942.

Coronavirus disease (COVID-19) pandemic. (2020). Retrieved from https://www.euro.who.int/en/health-topics/health-emergencies/coronavirus-covid-19#:~:text=WHO%20announced%20COVID%2D19,on%2011%20March%202020.

Davis, J. R., Wilson, S., Brock-Martin, A., Glover, S., & Svendsen, E. R. (2010). The impact of disasters on populations with health and health care disparities. Disaster Med Public Health Prep, 4(1), 30–38. doi:10.1017/s1935789300002391

De Sousa, A., Mohandas, E., & Javed, A. (2020). Psychological interventions during COVID-19: Challenges for low and middle income countries. Asian Journal of Psychiatry, 102128.

Degenhardt, L., Glantz, M., Evans-Lacko, S., Sadikova, E., Sampson, N., Thornicroft, G., … Helena Andrade, L. (2017). Estimating treatment coverage for people with substance use disorders: an analysis of data from the World Mental Health Surveys. World Psychiatry, 16(3), 299–307.

Dolan, A. H., & Walker, I. J. (2006). Understanding vulnerability of coastal communities to climate change related risks. Journal of Coastal Research, 1316–1323.

Dorahy, M. J., Rowlands, A., Renouf, C., Hanna, D., Britt, E., & Carter, J. D. (2015). Impact of average household income and damage exposure on post-earthquake distress and functioning: A community study following the F ebruary 2011 C hristchurch earthquake. British Journal of Psychology, 106(3), 526–543.

Dunlop, A., Lokuge, B., Masters, D., Sequeira, M., Saul, P., Dunlop, G., … Haber, P. (2020). Challenges in maintaining treatment services for people who use drugs during the COVID-19 pandemic. Harm Reduction Journal, 17, 1–7.

Dutra, S. J., Marx, B. P., McGlinchey, R., DeGutis, J., & Esterman, M. (2018). Reward ameliorates posttraumatic stress disorder-related impairment in sustained attention. Chronic Stress, 2, 2470547018812400.

European Monitoring Centre for Drugs and Drug Addiction and Europol (2020), EU Drug Markets: Impact of COVID-19, Publications Office of the European Union, Luxembourg. Accessed September 2020 at: https://www.emcdda.europa.eu/publications/ad-hoc/covid-19-and-drugs-drug-supply-via-darknet-markets_en

European Monitoring Centre for Drugs and Drug Addiction (May, 2020). Impact of COVID-19 on drug services and help-seeking in Europe. Accessed September 2020 at: https://www.emcdda.europa.eu/publications/ad-hoc/impact-of-covid-19-on-drug-services-and-help-seeking-in-europe_en

Farhoudian, A., Baldacchino, A., Clark, N., Gerra, G., Ekhtiari, H., Dom, G., … Demasi, M. (2020). COVID-19 and substance use disorders: recommendations to a comprehensive healthcare response. An international society of addiction medicine (ISAM) practice and policy interest group position paper. Autonomic Neuroscience: Basic & Clinical, 11(2), 129–146.

Farhoudian, A., Radfar, S. R., Mohaddes Ardabili, H., rafei, p., ebrahimy, m., Khojasteh zonoozi, A., … Ekhtiari, H. (2020). A global survey on changes in the supply, price and use of illicit drugs and alcohol, and related complications during the 2020 COVID-19 pandemic. medRxiv, 2020.2007.2016.20155341. doi:10.1101/2020.07.16.20155341

Fiorillo, A., & Gorwood, P. (2020). The consequences of the COVID-19 pandemic on mental health and implications for clinical practice. European Psychiatry, 63(1).

Forouzanfar, M. H., Afshin, A., Alexander, L. T., Anderson, H. R., Bhutta, Z. A., Biryukov, S., … Charlson, F. J. (2016). Global, regional, and national comparative risk assessment of 79 behavioural, environmental and occupational, and metabolic risks or clusters of risks, 1990–2015: a systematic analysis for the Global Burden of Disease Study 2015. The lancet, 388(10053), 1659–1724.

Frank, A., Fatke, B., Frank, W., Förstl, H., & Hölzle, P. (2020). Depression, dependence and prices of the COVID-19-Crisis. Brain, behavior, and immunity.

Han, H.-J., Kim, J.-H., Chung, S.-E., Park, J.-H., & Cheong, H.-K. (2018). Estimation of the national burden of disease and vulnerable population associated with natural disasters in Korea: heavy precipitation and typhoon. Journal of Korean Medical Science, 33(49).

Havakuk, O., Rezkalla, S. H., & Kloner, R. A. (2017). The cardiovascular effects of cocaine. Journal of the American College of Cardiology, 70(1), 101–113.

International Convention on the Protection of the Rights of All Migrant Workers and Members of Their Families. Retrieved from https://ohchr.org/EN/ProfessionalInterest/Pages/CMW.aspx

Khatri, U. G., & Perrone, J. (2020). Opioid Use Disorder and COVID-19: Crashing of the Crises. Journal of Addiction Medicine.

Knopf, A. (2020). Addiction telemedicine comes into its own with COVID-19. Alcoholism & Drug Abuse Weekly, 32(13), 5–6.

Lai, C.-C., Shih, T.-P., Ko, W.-C., Tang, H.-J., & Hsueh, P.-R. (2020). Severe acute respiratory syndrome coronavirus 2 (SARS-CoV-2) and corona virus disease-2019 (COVID-19): the epidemic and the challenges. International journal of antimicrobial agents, 105924.

Liu, C. H., & Doan, S. N. (2020). Psychosocial Stress Contagion in Children and Families During the COVID-19 Pandemic. Clinical Pediatrics, 0009922820927044.

Marsden, J., Darke, S., Hall, W., Hickman, M., Holmes, J., Humphreys, K., … West, R. (2020). Mitigating and learning from the impact of COVID-19 infection on addictive disorders. Addiction.

Masozera, M., Bailey, M., & Kerchner, C. (2007). Distribution of impacts of natural disasters across income groups: A case study of New Orleans. Ecological Economics, 63(2-3), 299–306.

Mellis AM, P. M., J Hulsey (In press). COVID-19-related treatment service disruptions among people with single-and poly-substance use concerns. J Substance Abuse Treatment.

Mongan, D., Galvin, B., Farragher, L., Dunne, M., & Nelson, M. (2020). Impact of COVID-19 on drug services in four countries.

Nagelhout, G. E., Hummel, K., de Goeij, M. C., de Vries, H., Kaner, E., & Lemmens, P. (2017). How economic recessions and unemployment affect illegal drug use: a systematic realist literature review. International Journal of Drug Policy, 44, 69–83.

Nations, U. (2020). World Drug Report 2020. Retrieved from

Nazari, S. S. H., Noroozi, M., Soori, H., Noroozi, A., Mehrabi, Y., Hajebi, A., … Mirzazadeh, A. (2016). The effect of on-site and outreach-based needle and syringe programs in people who inject drugs in Kermanshah, Iran. International Journal of Drug Policy, 27, 127–131. doi:https://doi.org/10.1016/j.drugpo.2015.10.011

Needle, R. H., Burrows, D., Friedman, S. R., Dorabjee, J., Touzé, G., Badrieva, L., … Latkin, C. (2005). Effectiveness of community-based outreach in preventing HIV/AIDS among injecting drug users. International Journal of Drug Policy, 16, 45–57. doi:https://doi.org/10.1016/j.drugpo.2005.02.009

Nobles, J., Martin, F., Dawson, S., Moran, P., & Savovic, J. (2020). The potential impact of COVID-19 on mental health outcomes and the implications for service solutions: Bristol, UK: National Institute for Health Research, University of Bristol.

O’Sullivan, T., & Bourgoin, M. (2010). Vulnerability in an influenza pandemic: Looking beyond medical risk. behaviour, 11, 16.

Onder, G., Rezza, G., & Brusaferro, S. (2020). Case-fatality rate and characteristics of patients dying in relation to COVID-19 in Italy. Jama, 323(18), 1775–1776.

Organization, W. H. (2010). Atlas on substance use (2010): resources for the prevention and treatment of substance use disorders: World Health Organization.

Peak, A., Rana, S., Maharjan, S. H., Jolley, D., & Crofts, N. (1995). Declining risk for HIV among injecting drug users in Kathmandu, Nepal: the impact of a harm-reduction programme. AIDS (London, England), 9(9), 1067–1070. doi:10.1097/00002030-199509000-00013

Peavy, K. M., Darnton, J., Grekin, P., Russo, M., Green, C. J. B., Merrill, J. O., … Tsui, J. I. (2020). Rapid Implementation of Service Delivery Changes to Mitigate COVID-19 and Maintain Access to Methadone Among Persons with and at High-Risk for HIV in an Opioid Treatment Program. AIDS and Behavior, 1.

Pfefferbaum, B., & North, C. S. (2020). Mental Health and the Covid-19 Pandemic. New England Journal of Medicine. doi:10.1056/NEJMp2008017

Phibbs, S., Kenney, C., Rivera-Munoz, G., & Huggins, T. J. (2018). The Inverse Response Law: Theory and Relevance to the Aftermath of Disasters. 15(5). doi:10.3390/ijerph15050916

Ren, S.-Y., Gao, R.-D., & Chen, Y.-L. (2020). Fear can be more harmful than the severe acute respiratory syndrome coronavirus 2 in controlling the corona virus disease 2019 epidemic. World journal of clinical cases, 8(4), 652.

Riezzo, I., Fiore, C., De Carlo, D., Pascale, N., Neri, M., Turillazzi, E., & Fineschi, V. (2012). Side effects of cocaine abuse: multiorgan toxicity and pathological consequences. Current medicinal chemistry, 19(33), 5624–5646.

Roncero, C., García-Ullán, L., de la Iglesia-Larrad, J. I., Martín, C., Andrés, P., Ojeda, A., González-Parra, D., Pérez, J., Fombellida, C., Álvarez-Navares, A., Benito, J. A., Dutil, V., Lorenzo, C., & Montejo, Á.L. (2020). The response of the mental health network of the Salamanca area to the COVID-19 pandemic: The role of the telemedicine. Psychiatry research,;291:113252. doi: 10.1016/j.psychres.2020.113252. Epub 2020 Jul 2.

Rowe, C., Santos, G.-M., Vittinghoff, E., Wheeler, E., Davidson, P., & Coffin, P. O. (2016). Neighborhood-level and spatial characteristics associated with lay naloxone reversal events and opioid overdose deaths. Journal of Urban Health, 93(1), 117–130.

Runkle, J. D., Brock-Martin, A., Karmaus, W., & Svendsen, E. R. (2012). Secondary surge capacity: a framework for understanding long-term access to primary care for medically vulnerable populations in disaster recovery. American journal of public health, 102(12), e24–e32. doi:10.2105/AJPH.2012.301027

Samuels, E. A., Clark, S. A., Wunsch, C., Keeler, L. A. J., Reddy, N., Vanjani, R., & Wightman, R. S. (2020). Innovation during COVID-19: Improving addiction treatment access. Journal of Addiction Medicine.

Samuels, E. A., Clark, S. A., Wunsch, C., Keeler, L. A. J., Reddy, N., Vanjani, R., & Wightman, R. S. (2020). Innovation During COVID-19: Improving Addiction Treatment Access. J Addict Med. doi:10.1097/adm.0000000000000685

Singer, M., Bulled, N., Ostrach, B., & Mendenhall, E. (2017). Syndemics and the biosocial conception of health. The Lancet, 389(10072), 941–950.

Smit, B., & Wandel, J. (2006). Adaptation, adaptive capacity and vulnerability. Global Environmental Change, 16(3), 282–292. doi:https://doi.org/10.1016/j.gloenvcha.2006.03.008

Sokol, R., Gupta, A., Powers, S., Hoffman, L., & Meza, J. (2020). Guidance for Treating Patients with Opioid Use Disorder (OUD) with Buprenorphine-Naloxone (B/N) in the COVID-19 Era via Telehealth: A Review of Previous Evidence, New COVID-19 OUD Treatment Guidelines, and a Case Report of their Application.

Spooner, C., & Hetherington, K. (2005). Social determinants of drug use: National Drug and Alcohol Research Centre, University of New South Wales ….

Starcke, K., & Brand, M. (2012). Decision making under stress: a selective review. Neuroscience & Biobehavioral Reviews, 36(4), 1228–1248.

Szabo, G., & Mandrekar, P. (2009). A recent perspective on alcohol, immunity, and host defense. Alcoholism: Clinical and Experimental Research, 33(2), 220–232.

Vasylyeva, T. I., Smyrnov, P., Strathdee, S., & Friedman, S.R. Challenges posed by COVID-19 to people who inject drugs and lessons from other outbreaks. Journal of the International AIDS Society, e25583.

Volkow, N. D. (2020). Collision of the COVID-19 and addiction epidemics: American College of Physicians.

Watt, G. (2002). The inverse care law today. Lancet, 360(9328), 252–254. doi:10.1016/s0140-6736(02)09466-7

World Economic Situation and Prospects 2019. (2019). Retrieved from https://www.un.org/development/desa/dpad/publication/world-economic-situation-and-prospects-2019/

Ying, S., Yang, S., & Jianming, F. (2020). Psychiatry hospital management facing COVID-19: from medical staff to patients. Brain Behav Immun, 10.

Zarrabian, S., & Hassani-Abharian, P. Covid-19 Pandemic and the Importance of Cognitive Rehabilitation. Basic and Clinical Neuroscience, 189–190.

